# Communication attributes modify the anxiety risk associated with social media addiction: a prospective diary method study

**DOI:** 10.1101/2022.11.30.22282943

**Authors:** Chenziheng Allen Weng, Jahshara Bulgin, Savannah Diaz, Jiafang Zhang, Runzi Tan, Le Li, Mari Armstrong-Hough

## Abstract

**Purpose:** Social media use in younger people has shown mixed associations with mental health. We hypothesized that communication types during social media use might alter the relationship between social media dependence and anxiety over time. We aimed to identify how four dimensions of communication influence the link between social media addiction (SMA) and anxiety.

**Methods:** We recruited a cohort of undergraduate students aged 18-26 to participate in daily surveys over two weeks using a diary method to assess daily social media use, SMA, anxiety symptoms, and the four dimensions of communication: Consumption, Broadness, Online Exclusivity, and Parasociality. Lagged logistic regression models with generalized estimating equations evaluated the influence of daily SMA and communication type on subsequent anxiety levels.

**Results:** Out of 79 participants, 1009 daily records were analyzed. SMA positively correlated with anxiety (Kendall rank correlation τ=.30). Interaction analysis indicated that levels of parasociality and consumption moderated the association between SMA components and anxiety outcomes. In young adults with high levels of consumption or parasociality, a 1-standard-deviation rise in SMA’s social conflict component led to an 11%-13% increase in next-day anxiety scores. This association was absent for those with low to moderate levels of parasociality and consumption.

**Discussion:** Elevated levels of passive consumption and one-sided interactions amplify the anxiety risk associated with social media dependence. Further longitudinal evidence can elucidate the connections between communication types, social media exposure, and anxiety, guiding the development of a model for healthy social media use.

**Implications and Contribution:** Problematic social media use among youth necessitates targeted interventions. This study demonstrates that passive consumption and one-sided social interactions significantly heighten the anxiety risks tied to social media addiction. The findings highlight the importance of considering communication types in developing interventions aimed at reducing social media-induced mental health issues.

**Highlights:**

- We examined four dimensions of social media communication via a daily diary method.
- Consumption and parasociality moderated the SMA-anxiety link, amplifying its effects.
- High consumption and parasociality predicted increased next-day anxiety scores.
- Communication attributes are important in understanding social media use

## 1 Introduction

The past two decades have seen a surge in social media use, with the percentage of American adults on these platforms jumping from 5% in 2005 to 72% by early 2021 (Hootsuite, 2021); young users aged 18-29 reported checking platforms like Instagram or Snapchat multiple times daily (Auxier & Anderson, 2021). Concurrently, the mental health effects of social media use, such as on loneliness (Teppers et al., 2014), stress management (Brand et al., 2016), attention span (Throuvala et al., 2019), and addictive behaviors (Moqbel & Kock, 2018; Shensa et al., 2017), have become increasingly scrutinized.

Social media addiction (SMA) is considered a behavioral addiction, characterized by preoccupation, salience, mood modification, tolerance, withdrawal, conflict, relapse, and persistent use despite adverse consequences, mirroring criteria for substance use disorders (Montag et al., 2024; Moretta et al., 2022). Evidence from longitudinal studies and meta-analyses have associated SMA with anxiety and depression (Huang, 2020; Keles et al., 2020). A nationally representative U.S. panel study from 2014 to 2016 observed a 9% increase in the odds of depression symptoms for young adults with problematic social media use (Shensa et al., 2017), while a multi-country survey linked intense SMA in school-aged adolescents with lower well-being across various domains (Boer et al., 2020). Thus, a better understanding of the mechanisms linking SMA to adverse mental health outcomes is essential for advancing theoretical models and guiding future research.

Although SMA itself has been well researched, previous work has been limited by an undifferentiated concept of social media activities that did not attend to differences between individuals in how or why they use social media. For example, a group of people primarily using the sites for making friends and chatting may experience different risk levels compared to another group using most of the time browsing content (Burke et al., 2010; Utz & Breuer, 2017). Scientific understanding remains limited regarding how different types of social media communication may influence the relationship between SMA and mental health outcomes, with few studies investigating whether SMA’s effects vary across communication types (Kuss & Griffiths, 2017; Marino et al., 2018).

To derive the SNS communication attributes defined by clear-defined, broader dimensions based on their shared characteristics and potential impacts on well-being, we built upon prior research linking specific communication types—such as targeted composed messages, active social interactions, and “one-click” activities like likes and shares—to mental health outcomes (Burke & Kraut, 2016; Manuoğlu & Uysal, 2020; Utz & Breuer, 2017). The present study examined SNS communication attributes across four distinct dimensions: 1. *Consumption*: creating content versus consuming content; 2. *Broadness*: whether information or messages sent or received were targeted/private versus broadcasted/public; 3. *Online Exclusivity*: whether connections or interactions took place exclusively on SNS versus in both SNS and other settings; 4. *Parasociality*: whether connections or interactions on SNS were one-way versus mutual. Detailed operational definitions and measurement procedures are described in the Methods section.

Previous studies have found communication types appear to be correlated with mental well-being (Burke & Kraut, 2016). For example, Burke and Kraut (2016) reported that receiving targeted, composed communication from strong-tie friends was associated with improvements in psychological well-being while viewing broadcasted messages and receiving one-click feedback were not (Burke & Kraut, 2016). In a survey of high school students, Frison and Eggermont (2016) found that the incidence of depressed mood was higher among those who used Facebook passively, defined as viewing others’ profiles and posts (Frison & Eggermont, 2016). This line of work is especially invaluable in putting novel, online communication in the context of established models of human needs for information exchange and social connections. Does it facilitate essential social efforts (e.g., facilitating belongingness, signaling relational investment, maintaining stock of friendships, and bolstering self-affirmation (Manuoğlu & Uysal, 2020; Toma & Hancock, 2013; Tsao et al., 2021))? Or does it perhaps amplify morbid psychosocial mechanisms (e.g., stimulating unhealthy social comparison, sensation seeking, ignoring real-life relationships, and causing decreased productivity (Faelens et al., 2021; Griffiths, 2013; Kircaburun & Griffiths, 2018; Leung, 2008))?” Collectively, this body of empirical evidence provides a compelling rationale for differentiating SNS communication based on contextual factors. However, a critical knowledge gap persists regarding how these distinct communication patterns potentially moderate the association between SMA and psychological outcomes - the primary objective of the present investigation.

This paper aims to investigate how specific online communication attributes moderate the link between SMA and anxiety. Building on prior work of Burke and Kraut (2016) and Shakya et al. (2017), we systemize four dimensions of communication – Consumption, Broadness, Online Exclusivity, and Parasociality – into coherent framework to assess their moderating roles. Employing a daily diary design to collect prospective data, this study captures day-to-day fluctuations in social media addiction (SMA) and anxiety. We expected to offer a temporal perspective to strengthen empirical research on communication types and to test the hypothesis that the effects of SMA on anxiety would vary by exposure to different types of social media communication.

## 2 Methods

### 2.1 Study Design

The study was designed to collect prospective behavioral data through daily diary method (Shiffman et al., 2008). Individual participants’ daily social media activities and dimensions of communication, measures of SMA, and anxiety levels were repeatedly captured and analyzed longitudinally across a 14-day period. Data collection procedures were designed to minimize recall bias, consistently record daily measures, and deliver daily surveys on mobile devices in the same context as participants used social media.

The study population included current full-time NYU undergraduate students (9 credits or above per semester) between 18-26 years old and currently residing in the United States. Potential participants were excluded if they were not regular users of major social networking site (SNS) applications. We defined SNS as any social media application that connects users via user-created individual profiles (Kuss & Griffiths, 2011). Examples of SNS applications/platforms given to participants at recruitments and during daily surveys include Facebook, Snapchat, WeChat, Instagram, Twitter, TikTok, Weibo, LinkedIn, Reddit, and Tumblr.

*Ethics approval and consent to participate*

Ethical approval for the study was obtained from the New York University Institutional Review Board (IRB-FY2020-4578). Informed consent to participate was obtained from all of the participants in the study.

#### 2.1.1 Study size

Through a review of previous diary method studies with a similar study population, we found that the range of study size and time points fall around 75-120 subjects and last from 5 to 21 days (Liu & Lou, 2019; Ohly et al., 2010). We selected 14 days to balance variance against risk of response fatigue.

We conducted a standard statistical power analysis under Repeated Measures ANOVA for within-group factors. To detect an effect size of 0.06, the number of measurements at 14, and the power set at 0.80, the required sample size is 90. G*Power 3.1 (Faul et al., 2007) was used for the statistical power analysis and study size determination.

### 2.2 Procedures

Data were collected from March 2021 to May 2021. Recruitment emails were distributed to undergraduates at New York University through the institutional email system. The research team sent out three rounds of recruitment emails to reach the predetermined sample size of 90.

Completion of the study involved three phases: an orientation session through a prerecorded tutorial video and web page, a baseline questionnaire, and 14 consecutive days of diary records. During the study period, participants received an email each day at 8pm directing them to the survey. Participants were required to finish the diary survey within 3 hours to stay in the study. Three reminders were sent via email. In cases of extenuating circumstances, participants may make up a missing diary survey until 12:00 pm on the following day up to three times by notifying their administrative investigator. Design, distribution, and data collection were conducted using Qualtrics XM.

At the beginning of the daily survey, participants completed calibration questions to systematically review their social media usage. These questions consisted of (1) a selection of all SNS applications they had used since the last check-in, where the ten most popular SNS platforms were listed and a free text entry was provided for other platforms; (2) a number entry of an estimation of the total number of times and the total time spent in checking the SNSs, broken down into evening, morning, and afternoon. The main daily-level questionnaire was then administrated block by block. The survey did not allow skipped responses.

Participants received USD $25 compensation for completing 14 consecutive diary days and $10 for completing at least 7 consecutive days.

### 2.3 Constructs and Measures

#### 2.3.1 Time-independent measures: baseline characteristics

Demographic variables including gender, age, and race/ethnicity were measured by self-report in the baseline survey. Measures of four control variables were collected at baseline: history of diagnosis with a depression disorder, history of diagnosis with an anxiety disorder, recent experience of a major exam or major life event (e.g., marriage, breakup, losing a job, or the death of a loved one), and COVID-19-related adjustment disorder. *COVID-19-related adjustment disorder* was measured by an adapted International Adjustment Disorder Questionnaire (IADQ) (Shevlin et al., 2020), as shown in Supplementary Textbox 1.

#### 2.3.2 Time-dependent measures: Daily-level entries

*Anxiety* is the primary outcome of interest and was measured by the Generalized Anxiety Disorder scale, known as the GAD-7 (Löwe et al., 2008). The scale consists of seven questions with a 0-3 measured Likert-type scale, which were adapted for daily measures. The total score is the sum of each question. A score >= 10 was considered to indicate anxiety symptoms, with a score of 10-14 indicating moderate symptoms and a score >= 15 indicating severe symptoms.

*SNS communication type:* we defined *Consumption* as the proportion of time spent viewing content (+), compared to the proportion spent composing content (–). We defined *Broadness* as, among messages sent and received, what proportion were broadcasted to a broad audience (+), compared to the proportion that was targeted at a particular person (–). *Online exclusivity* was defined as, among connections fostered on social media sites, what proportion of relationships existed online-only (+, e.g., net friends, celebrities), compared to the proportion existing offline as well (–, e.g., classmates, family members). <u>Parasociality</u> was defined as, among connections they made on social media sites, what proportion was primarily one-way (+), while what proportion was primarily mutual (–). Participants were asked to assess their communications on SNS since the last study contact on a bipolar scale from 1 to 10 for each dimension. The full scale is shown in Textbox 1.

*Social media addiction* was assessed by a set of 12 items adapted from the Bergen Facebook Addiction Scale (Andreassen et al., 2012), as shown in Supplementary Textbox 2. Each item represented one of four components of addiction: salience, mood modification, social conflict, and withdrawal. Each addiction component has three questions. Participants responded on a Likert-type scale with anchors from Rarely (0) to All the time (3). While the original items specified “Facebook,” we substituted “Facebook” with “social networking sites”. The resulting composite scale ranged from 0 to 36 and served as a primary independent variable in our model. The raw alpha and the standardized alpha were 0.79 and 0.80, respectively.

### 2.4 Statistical analysis

Intraclass correlations (ICC) for dimensions of communication and SMA were computed to describe the consistency of repeated daily-level measures of the same variable within individual participants over the study period. Generalized estimating modeling (GEE) was used to estimate how SMA dimensions and communication types predict changes in anxiety. We employed a lagged logistic regression framework where predictors, including dimensions of social media addiction (SMA) and communication types, as well as the lagged outcome (anxiety symptoms), were measured on the previous day (*t* – 1. This approach ensures temporal alignment, allowing us to model how behaviors and exposures on one day influence anxiety levels on the following day (i;). The inclusion of a lagged dependent variable (*Anxiety_t_*_-1_) accounts for temporal autocorrelation, providing more robust estimates of the predictors effects.

As a centering procedure, all predictors and outcomes were standardized to facilitate interpretation, with coefficients representing the change in the likelihood of next-day anxiety per 1-SD increase in predictors from the previous day. Additionally, the outcome variable was transformed to a relative scale so that regression coefficients represent the percent change in the anxiety outcome by each unit of 1-standard-deviation increase in independent variables. Further stratified analysis applied the final multivariable model on the tertiles of each communication dimension.

A significance level of 0.10 was chosen to enhance the sensitivity of the analysis, enabling the detection of potentially meaningful associations while maintaining an acceptable balance with Type I error risk. R software (version 4.3.0; R Foundation for Statistical Computing), including the *gee* package (version 4.13-25).

## 3 Results

### 3.1 Sample Characteristics

Among 94 participants who finished the baseline survey, 79 (84%) participants finished at least 3 daily (survey) records and were included in the analysis, resulting in 1009 total records analyzed (Table 1). On average, participants completed 13 daily records (median: 14).

**Table 1.**
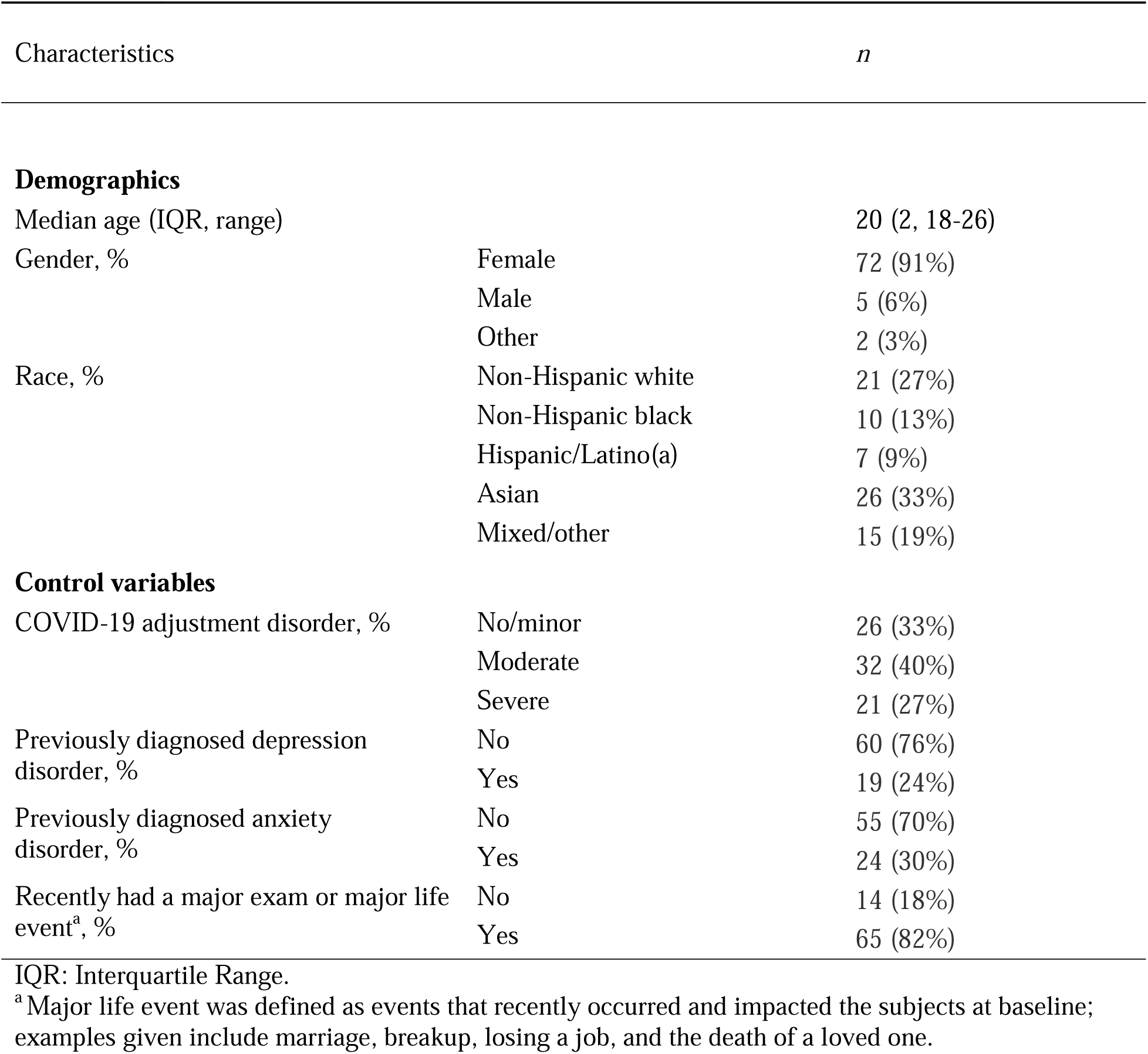
Study sample characteristics (*n*=79)

The median age was 20 years (range: 18–26), and the majority were women (91%). Over two-thirds (67%) reported moderate or severe levels of COVID-19 adjustment disorder, with 27% experiencing severe levels. Additionally, 24% and 30% reported previously diagnosed depression and anxiety disorders, respectively.

### 3.2 Distribution of daily-level measures

Distributions of the four communication dimensions are shown in Figure 1, and summary statistics of daily-level measures are presented in Supplementary Table 3. Consumption showed the largest within-subject variance (ICC=0.40), followed by broadness (ICC=0.58), online exclusivity (ICC=0.54), and parasociality (ICC=0.53).

**Figure 1.**
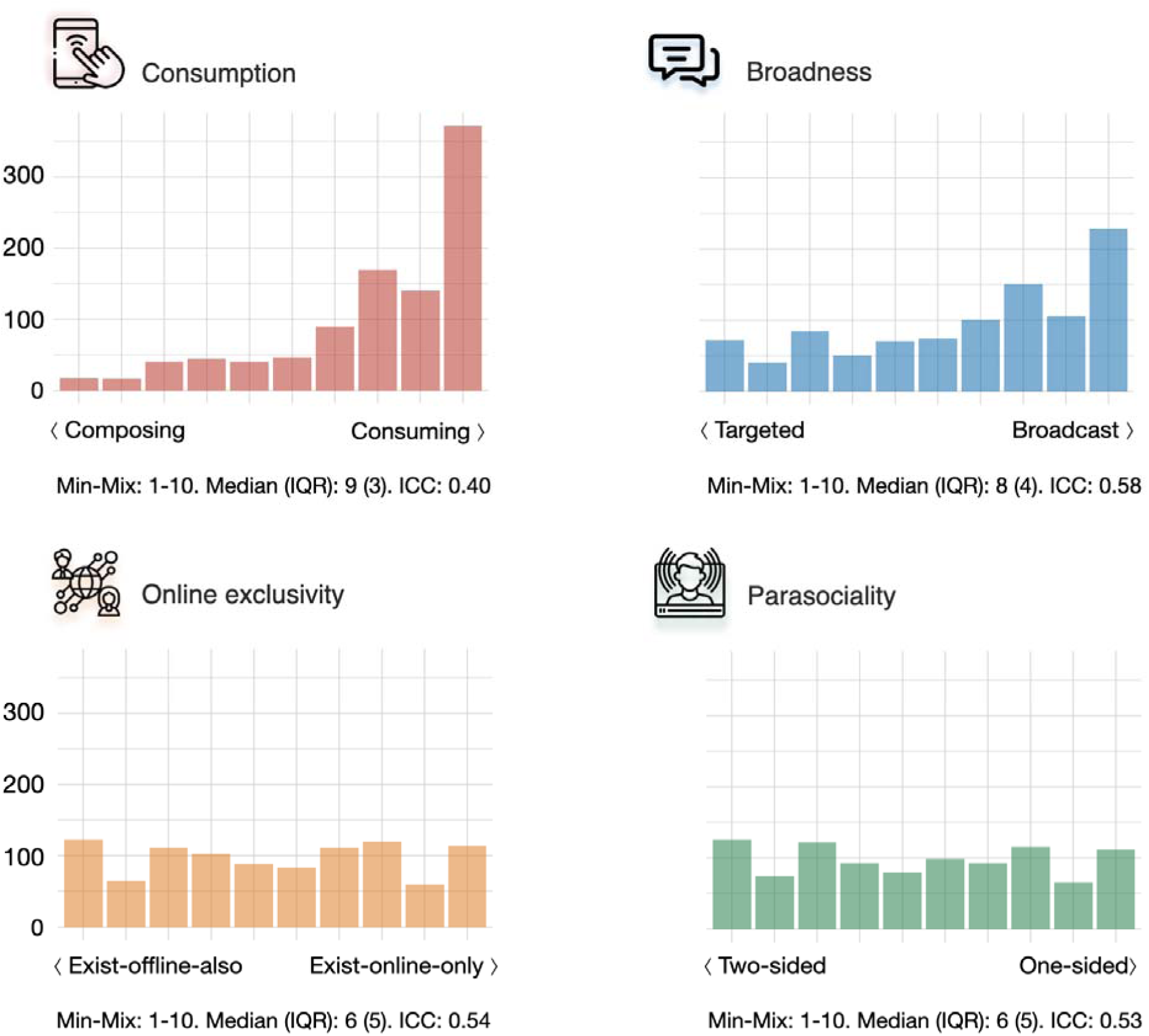
Distribution of the four dimensions of communication type of the daily survey records (n=1009). Axis labels show the visual analog of the self-reported relative proportion of time on the type of communication exchanged with a range of one to ten. ICC was measured in two-way random effects models. A higher ICC score indicates that the values from individual participants were more consistent over time. IQR: Interquartile range. ICC: Intraclass correlation.

All four SMA components (salience, mood modification, social conflict, and withdrawal) were significantly correlated with anxiety scores (P<0.001), with correlation coefficients ranging from 0.08 to 0.35. Salience and withdrawal showed moderate positive relationships with anxiety, while mood modification and social conflict demonstrated smaller and notable relationships, respectively. Communication dimensions were not significantly associated with anxiety scores in bivariate analysis.

### 3.3 Multivariable regression

Table 2 presents the population-averaged effect from GEE models, in which day-to-day change in daily anxiety score was regressed on previous-day SMA dimensions, previous-day communication characteristics, and their interactions. All models use a lagged dependent variable and were adjusted for age, gender, race, COVID-19 adjustment disorder, major life events or exams, and previously diagnosed anxiety or depression disorder.

**Table 2.**
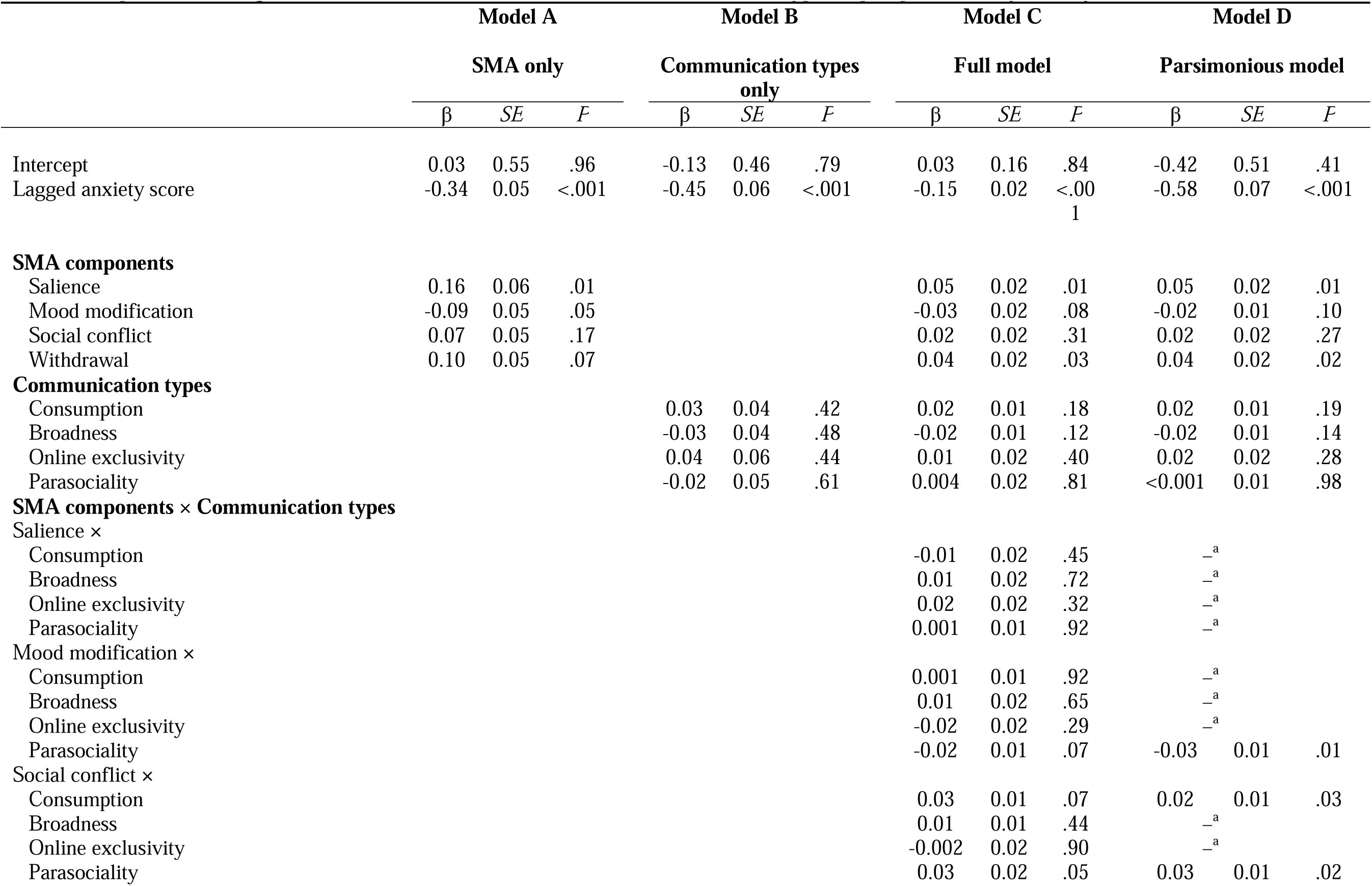
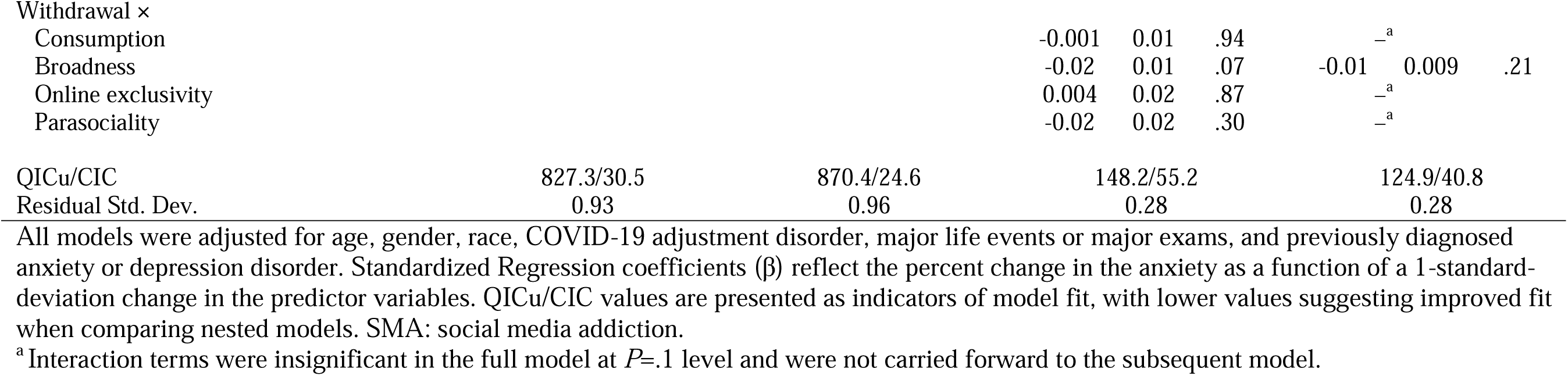
Population-averaged effect of social media addiction and communication type on prospective daily anxiety score (*n*=1009)

Previous-day anxiety score was negatively associated with next-day anxiety level in all models (Table 2, Model D, β=-0.58, *P*<0.001). Without accounting for attributes of communication, the salience component of SMA was associated with increased anxiety level (Table 2, Model A, β=0.16, *P*=0.01). The effect of withdrawal was also positively associated with anxiety score (β=0.10, *P*=0.07). Mood modification was associated with decreased anxiety level (β=-0.09, *P*=0.05). The effect of social conflict was not statistically significant (*P*>0.1). In Model B, communication characteristics are added to the model. None of the communication dimensions were independently associated with anxiety level. The full model (Model C) includes SMA components, communication characteristics, and the interaction terms of all pairwise combinations (4×4) of these dimensions. The main effects of SMA components and communication characteristics are similar to those in the first two crude models. Four significant interaction effects stand out: parasociality negatively moderated the effect of mood modification; consumption and parasociality positively moderated the effect of social conflict; broadness negatively moderated the effect of withdrawal.

In the parsimonious model (Model D), parasociality negatively moderated the effect of mood modification (β =-0.03, *P*=0.01); consumption and parasociality positively moderated the effect of social conflict (β=0.02, *P*=0.02; and β=0.03, *P*=0.02, respectively). That is, for every 1-standard-deviation increase in consumption, the effect of social conflict on next-day anxiety increased by 2% (95% CI 0.04%-4%). For every 1-standard-deviation increment in parasociality, the effect of social conflict on next-day anxiety increased by 3% (95% CI 1%-5%). The interaction of broadness and withdrawal was not statistically significant.

Model fit improved significantly in the full and parsimonious models accounting for the interactions between SMA components and communication types.

### 3.4 Stratified by communication types

Stratified analysis further examined the effects of SMA based on the levels of the moderating dimensions of communication (Figure 2). For the three pairs of significant interactions identified in the multivariable analysis, the models were rerun on the tertiles of their corresponding dimension of communication.

**Figure 2.**
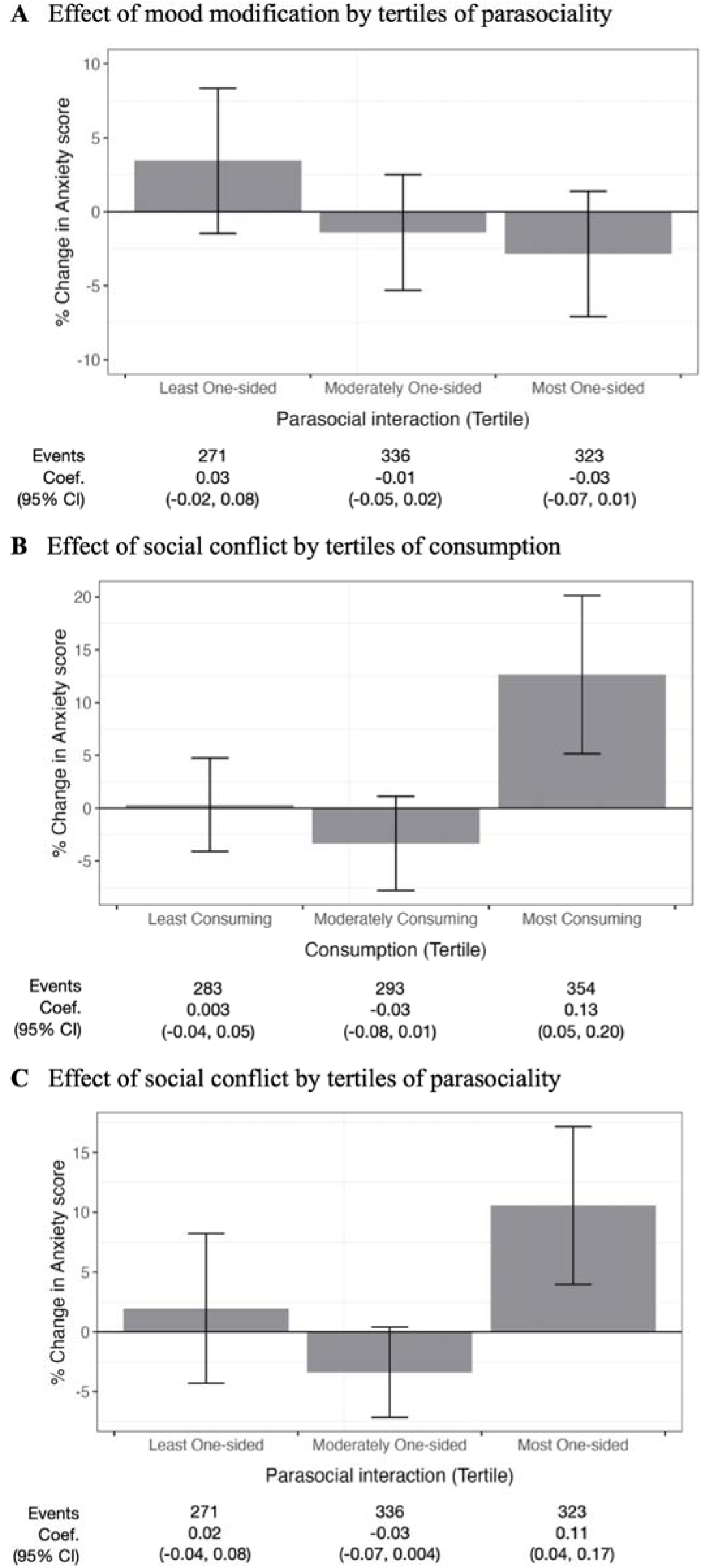
Stratified analysis of the effects of SMA components on anxiety based on tertiles of the dimensions of communication type. Coefficients indicate percentage changes in the outcome as a function of a 1-standard-deviation increase in the SMA component. SMA: social media addiction.

For participants with high consumption, a 1-standard-deviation increase in social conflict was associated with a 13% increase in next-day anxiety (95% CI 5%–20%), while no significant effects were observed for low-consumption groups (*P*>0.1). For parasociality, high levels amplified the effect of social conflict on anxiety, leading to an 11% increase in next-day anxiety (95% CI 4%–17%), while effects in low-parasociality groups were not significant (*P*>0.1). No significant effects were observed for interaction between parasociality and mood modification in stratified analyses (*P*>0.1).

## 4 Discussion

In this analysis of the moderating role of social media use type in the association between SMA and anxiety among young adults, we found that not all styles of social media use carry the same risks of heightened anxiety. Young adults whose social media interactions were characterized by consumption rather than composition experienced significant increases in SMA-induced anxiety. Similarly, young adults with primarily parasociality experienced significant increases in SMA-induced anxiety, an effect not observed in those with low or moderate interaction levels.

We also observed a pronounced effect of social conflict in the interactions between SMA and communication types. The results indicate that SMA problems may disrupt social obligations and focus on tasks, making individuals more susceptible to interpersonal tensions and productivity-related conflicts. This aligns with prior studies pointing out that behaviors characterizing social conflict are the most observable among SMA components (Griffiths, 2005; Shakya & Christakis, 2017). Recognizing this, it’s crucial to investigate further how different facets of social media use can either exacerbate or alleviate these conflicts, especially since such conflicts have profound implications for mental health and daily functioning. More comprehensive research on social conflict induced by SMA will be essential to inform interventions aimed at reducing the adverse effects of problematic social media use.

A core finding was the negative impact of excessive consumption as opposed to composition, or what is often termed “passive use”, on anxiety. This is consistent with Frison’s study which linked passive use of social media with depressive moods in adolescent girls, ascribed largely to upward social comparison (Frison & Eggermont, 2016). Conversely, active use, encompassing activities like posting photos and sending personal messages, was found beneficial due to the perception of increased online social support. Our findings reaffirm the potential dangers of passive consumption, particularly given the personalized content and highly self-relevance information prevalent on social media platforms (Lindström et al., 2021; Scholz et al., 2023). These patterns of consumption have been linked to sensation seeking (Bai et al., 2021; Leung, 2008), attentional deficits (Uncapher & Wagner, 2018), and even structural brain changes among young adults (Uncapher & Wagner, 2018). Future research should delve deeper into the mechanisms to explain how consumption exacerbates SMA-related anxiety.

Our research also underscored role of parasociality in exacerbating SMA-related anxiety. Engaging in parasociality, which are devoid of genuine relational feedback (Burke & Kraut, 2016; Throuvala et al., 2019; Utz & Breuer, 2017), can detract from time spent on meaningful, supportive relationships (Giles, 2020). Interestingly, increased parasociality was observed to have an almost immediate impact on next-day anxiety, suggesting potential mechanisms beyond just impairing other social interactions. While some argue that moderate levels of parasociality can foster feelings of community and identity exploration among young adults (Hoffner & Bond, 2022), our findings emphasize the importance of understanding and balancing such interactions to mitigate potential negative effects on mental well-being.

Our findings contribute to the growing body of literature emphasizing the cognitive implications of social media behaviors and their parallels in real-world social interactions (see a review by Meshi et al. (2015)). Adopting a “addictive” perspective of social media use, despite ongoing debates about its operational and theoretical boundaries (Montag et al., 2024; Moretta et al., 2022), provides a structured framework for the application of validated addiction frameworks to study SMA’s behavioral and psychological mechanisms. It’s imperative to understand that communication types on social media platforms are often intertwined. While beyond the scope of the current study, a more comprehensive understanding of how communication dimensions interact is needed. Further research to examine these interrelations could ultimately inform interventions to promote healthier online communication.

## Limitations

This study’s conclusions are constrained by its dependence on participants’ self-reporting of their social media use, which introduces variability in the interpretation of activity types and hinders direct comparisons across subjects. Furthermore, reported proportions of communication types, such as “consumption,” may not accurately reflect actual time spent, complicating the assessment of exposure’s impact. We employed calibration questions to standardize activity ratings, required at the beginning of the daily survey to allow participants to break down the time, sessions, and platforms on social media and help them preserve an objective review and estimation. Our sensitivity analysis, which included converting self-reported data into actual usage time, confirmed the consistency of our results.

Secondly, the predominance of female participants in our sample suggests a need for subsequent research to incorporate a more balanced gender representation. Moreover, the timing of the study—conducted during the initial COVID-19 outbreak—might have influenced the generalizability of the findings, given the potential for heightened stress levels among participants. However, we controlled for gender and COVID-19-related distress in our regression and stratified analyses.

Notwithstanding these limitations, the study’s methodological strengths lie in its use of an ecological momentary assessment framework coupled with prospective design in regression estimation. This study design enhances the ecological validity of our findings and decreases retrospective bias, providing a reliable record of subjective experiences, anxiety trends, and the immediate effects of social media interaction.

## Conclusions

Using a daily diary design, we found that two dimensions of social media communication significantly worsened the risk of anxiety associated with SMA among young adults. Young adults with high levels of passive social media use as defined by consumption and parasociality experienced significantly higher risk of next-day anxiety. Future research should systematically examine how the nature of social media communication—rather than just its quantity—shapes anxiety risk. Deepening understanding of the mechanisms linking problematic social media use, communication type, and anxiety can inform interventions to promote healthier use of social media among young adults.

## Data Availability

All data produced in the present study are available upon reasonable request to the authors.

## Acknowledgments

The recruitment procedure of this study was greatly aided by Julie Avina, Ed.D., who lent her hands to the then-student co-authors of this study. The project was funded by NYU Internal Faculty Funding.

## CRediT authorship contribution statement

CAW: Conceptualization, Data curation, Formal Analysis, Methodology, Project administration, Writing – original draft. JB: Data curation, Methodology, Investigation, Writing – original draft. SD: Project administration, Investigation, Resources, Validation. JZ: Formal Analysis, Investigation, Software. RT: Conceptualization, Investigation, Software. LL: Investigation, Methodology. MAH: Validation, Writing – review & editing, Resources.

**Funding sources**

The study process required minimal funding from NYU internal faculty, and the funding source was not involved in the research process.

### Abbreviations

SMA: social media addiction
SNS: Social networking site

### Textbox 1. SNS communication type scale. SNS: social networking sites.

**Scoring:** Scored from 1 to 10 to indicate the proportion of time. Participants rate their daily SNS activities in terms of each communication type as relative deciles that add up to 100% of total activity time.

**Item 1 – Consumption**

Instructions: Generating content vs. Viewing content

**Item 2 – Broadness**

Instructions: The content you interact (send and receive) with are: Targeted at the recipient vs. Broadcast to many recipients

**Item 3 – Online exclusivity (Shown as “Social connections”)**

Instructions: The connections you made on SNS exist offline also (fellow students, dates, parents, etc.) vs. exist only online (celebrities, influencers, idols, etc.)

**Item 4 – Parasociality (Shown as “Direction of relationships”)**

Instructions: The connections you made on SNS were primarily one-way (e.g., following an influencer) vs. primarily mutual (e.g., interacting in a two-way conversation.)

### Supplementary Textbox 1. Measure of COVID-19-Related Adjustment Disorder.

Participants identified their most significant stressor since the onset of the pandemic in March 2020 from a list of ten options, including “Overwhelmed by information,” “Disrupted ability to work or learn,” “Death of a loved one,” “Moving,” “Reduced/suspended social life,” “Financial difficulties,” “Family conflict,” “Serious illness,” “Unemployment,” and “Illness/care of a loved one.” This selected stressor was used in the International Adjustment Disorder Questionnaire (IADQ) main scale, which consists of six items assessing difficulty in adaptation (e.g., “I find it difficult to relax and feel calm after [Overwhelmed by information]”) and functional impairment (e.g., “Has [Overwhelmed by information] affected your relationships or social life?”).

Participants rated these items on a Likert scale from 0 (No) to 3 (Extreme). A score of 10 or higher indicated moderate adjustment disorder, while a score of 14 or higher indicated severe adjustment disorder.

### Supplementary Textbox 2. Social media addiction scale. Dimensions of the items: Salience (s), Mood modification (m), Social conflict (c), Withdrawal (w). SNS: social networking sites.

**Scoring:** Scored 1 (Rarely), 2 (Sometimes) 3 (Over half of the day) 4 (All the time) for items. Higher scores indicate more addictive symptoms. SNS: social networking sites.

**Scale items:**

While I study/work, my mind remains on the social networking sites. (s) The sites help to lift my mood. (m)

I find it difficult to switch off while I am on the sites. (s) I feel like going to the sites whenever I am upset. (m)

I check my SNS accounts before starting any task or activity. (s) I feel at ease when I am on the sites. (m)

I prefer the excitement of the sites over being with my close friends. (c) I find myself anticipating when I will go on the sites again. (w)

I’ve given less priority to hobbies, leisure activities, and exercise because of time spent on the sites. (c)

I’ve become restless when I do not have time to log in to the sites. (w)

I feel defensive or secretive if someone bothers me while on the sites. (c) I feel frustrated when I cannot use the sites. (w)

**Supplementary Table 1.** Distribution, intraclass correlations, and association measures of daily-level social media behavioral variables. Variables are the aggregates from the 14-day follow-up

| Variables | Min | Max | Median (IQR) | ICC | Corr. With Anxiety Score |
| --- | --- | --- | --- | --- | --- |
| **Social media communication dimensions** |  |  |  |  |  |
| Consumption | 1 | 10 | 9 (3) | 0.40 | <0.01<sup>a</sup> |
| Broadness | 1 | 10 | 8 (4) | 0.58 | -0.01<sup>a</sup> |
| Online exclusivity | 1 | 10 | 6 (5) | 0.54 | <0.01<sup>a</sup> |
| Parasociality | 1 | 10 | 6 (5) | 0.53 | 0.02<sup>a</sup> |
| **Social media addiction (SMA)** |  |  |  |  |  |
| Salience | 3 | 12 | 6 (3) | 0.65 | 0.28<sup>b</sup> |
| Mood modification | 3 | 12 | 6 (2) | 0.65 | 0.08<sup>b</sup> |
| Social conflict | 3 | 12 | 4 (2) | 0.71 | 0.35<sup>b</sup> |
| Withdrawal | 3 | 12 | 4 (2) | 0.74 | 0.33<sup>b</sup> |
| Sum score | 12 | 48 | 20 (8) | 0.75 | 0.30<sup>b</sup> |
| **Anxiety score** | 7 | 28 | 12 (8) | 0.65 | / |

Note: Scores were measured by a GAD-7 anxiety scale adapted for daily measures. ICC was measured in two-way random-effect models. A higher ICC score indicates that the values from individual participants were more consistent over time. Correlation coefficients were measured by Kendall's rank correlation $\tau$ . IQR = Interquartile range. ICC = Intraclass correlation coefficient.
<sup>a</sup> $P>.1$
<sup>b</sup> $P<.001$

